# Associations of Outdoor Air Pollution With Cause-specific Mortality in Metastatic Prostate Cancer: A cohort study

**DOI:** 10.64898/2026.09.29.26364034

**Authors:** Andrea Cosenza, David-Dan Nguyen, Zhiyu Qian, Vincenzo Striano, Leonard Appleman, Francesco Montorsi, Alberto Briganti, Giorgio Gandaglia, Benjamin J. Davies, Bruce L. Jacobs, Quoc-Dien Trinh, Hari S. Iyer

## Abstract

**Background:** Cardiovascular disease is an important competing cause of death in men with metastatic prostate cancer, and ambient air pollution may increase cardiovascular vulnerability. We evaluated associations of fine particulate matter (PM2.5) and nitrogen dioxide (NO2) with all-cause, cardiovascular, and prostate cancer-specific mortality, including joint associations with androgen deprivation therapy (ADT).

**Methods:** We analyzed 33,384 men from 11 US state registries diagnosed with metastatic prostate cancer during 2000–2015 and followed through 2018. Residential PM2.5 and NO2 concentrations at diagnosis were assigned using 1-km exposure models. Multivariable Cox models estimated all-cause mortality, and inverse-probability-weighted Cox models assessed cause-specific mortality.

**Results:** Each interquartile-range increase in PM2.5 was associated with higher cardiovascular mortality (HR 1.04, 95% CI 1.02–1.07; p-trend=0.008). Cardiovascular mortality was also higher in the fourth and fifth PM2.5 quintiles versus the first (HR 1.18 and 1.21, respectively). Higher NO2 was associated with prostate cancer-specific mortality in the highest quintile (HR 1.07, 95% CI 1.01–1.13). In joint analyses, ADT-treated men with high versus low PM2.5 exposure had higher cardiovascular mortality (HR 1.19, 95% CI 1.05–1.34), without clear evidence of effect modification (p-interaction=0.051).

**Conclusions:** In men with metastatic prostate cancer, higher PM2.5 exposure was associated with cardiovascular mortality, particularly among ADT-treated patients, supporting further study of environmental exposures and systemic therapy in cardiooncology.

**What this study adds:** Men with metastatic prostate cancer receiving androgen deprivation therapy may be particularly vulnerable to cardiovascular complications. In this large population-based cohort, higher residential PM2.5 exposure was associated with increased cardiovascular mortality, including among men receiving ADT. Although we found no clear evidence that ADT modified the effect of PM2.5, the joint exposure pattern suggests that environmental air pollution may contribute to cardiovascular vulnerability in patients already exposed to cardiometabolic effects of systemic therapy. These findings extend environmental epidemiology to a clinically high-risk cancer population.

## Introduction

Prostate cancer (PCa) is among the most common malignancies in men and remains a leading cause of cancer-related mortality, particularly among patients with advanced or metastatic disease.^1^ In the metastatic setting, cardiovascular disease (CVD) represents an important competing cause of death and is among the leading causes of mortality.^2,3^ This issue is clinically relevant because androgen deprivation therapy (ADT), the standard treatment for metastatic disease, is associated with adverse cardiometabolic effects, including metabolic dysfunction, changes in body composition, and increased risk of cardiovascular events.^4,5^

In this context, identifying modifiable environmental factors that may exacerbate cardiovascular vulnerability among men with metastatic PCa is important for clinical management and risk stratification.^6^ Fine particulate matter air pollution (PM2.5) is the leading environmental cause of morbidity and mortality globally, contributing to 13% of deaths and 8% of disability-adjusted life-years each year.^6,7^ Long-term exposure to PM2.5 increases risk of cardiovascular disease (CVD) through multiple biological pathways including inflammation, oxidative stress, which can lead to hypertension and dyslipidemia that ultimately drive CVD.^8–12^ In addition, the increasing intensity and severity of wildfires has contributed to more individuals being exposed to unsafe levels of air pollution in recent years.^13,14^ These adverse cardiovascular effects may be particularly significant for patients with metastatic cancer due to comorbidities and side effects of treatment.^15^

Emerging evidence further suggests that air pollution may adversely affect cardiovascular health in patients receiving potentially cardiotoxic cancer therapies.^16^ Prior population-based work in men with prostate cancer has suggested that specific PM2.5 components are associated with higher all-cause and cardiovascular mortality, supporting the relevance of air pollution to non-cancer outcomes in this population.^10^ Whether this association extends to men with metastatic PCa, particularly those exposed to ADT, remains unclear. It is also unknown whether air pollution has a stronger impact in this clinically vulnerable population than in broader prostate cancer populations.

Therefore, we evaluated associations of PM2.5 and nitrogen dioxide (NO2) exposure with cardiovascular, all-cause, and prostate cancer-specific mortality among men with metastatic prostate cancer. We also examined joint ADT-pollution exposure categories to assess whether higher pollutant exposure was associated with mortality among treated patients. Our goal was to clarify whether environmental exposure could help inform cardiovascular risk stratification in this clinically vulnerable population.

## Methods

### Data source and study population

We conducted a population-based cohort study using data from the Multilevel Epidemiology Tumor Registry for Oncology (METRO), a multistate registry-based cohort including men diagnosed with prostate cancer between 2000 and 2015. States (California, Georgia, Hawaii, Louisiana, Massachusetts, Michigan (Detroit metropolitan area), New Jersey, New Mexico, Ohio, Pennsylvania, Utah, and Washington (Seattle/Puget Sound metropolitan area) were selected to capture variation in health services and environmental exposures related to PCa in different parts of the US.^17,18^ Registry data included demographic, clinical, treatment, residential, and mortality information, with follow-up through death, censoring on December 31, 2018, or 10 years from diagnosis. Geomasked residential addresses were obtained for participants, applying a jittering procedure within 250m in urban areas and 400m in rural areas to protect privacy.^17^ Further details about the METRO cohort have been published previously.^10^

The analytic cohort was restricted to men with distant-stage prostate cancer at diagnosis (n=44,609). We excluded men who were not aged 40-99 years (n=64), those who lacked pollution linkages (n=9236), and patients diagnosed with autopsy (with follow-up of 1 month or less) (n=1925) were excluded. None of the Hawaiian participants were retained, leading to a sample across 11 US states. Follow-up time was measured in months from diagnosis. The Dana-Farber Cancer Institute and Rutgers University, The State University institutional review boards approved this study and determined that, because existing data sources were used, no written consent was required for participation in the study. Further state institutional review board approval from California, Massachusetts, New Jersey, New Mexico and Utah was required and granted.

### Exposure assessment

The primary environmental exposures were predicted outdoor residential air pollutant values derived from ensemble models taking multiple geospatial meteorological, built and contextual environment, and source data and validated using air pollution monitors from government and academic sources produced by Di et al. with methodology described elsewhere.^19,20^ The models displayed 10-fold cross-validation R2 values of 0.89 for annual predictions of PM2.5 and 0.79 for NO2, indicating robust predictive accuracy, and were predicted over 1km grids for the contiguous United States from 2000-2016. We linked annual averaged residential PM2.5 and NO2 exposures to participants’ geomasked residential address during year of diagnosis. PM2.5 and NO2 were evaluated both as continuous variables scaled per interquartile range (IQR) increase and as quintiles of exposure, with the lowest quintile used as the reference group with an ordinal p-for-trend. For joint treatment-pollution analyses, PM2.5 and NO2 were dichotomized at the cohort median (PM2.5 = 10.3 µg/m^3^, NO2 = 24.1 ppb) into low and high exposure. Androgen deprivation therapy (ADT) exposure was defined using registry-based hormone therapy information. Four-category joint exposure variables were then created separately for PM2.5 and NO2: ADT with low exposure, ADT with high exposure, no ADT with low exposure, and no ADT with high exposure. The reference category for these models was ADT with low pollutant exposure.

### Outcomes

The primary outcome was cardiovascular disease (CVD)-specific mortality, defined using registry-recorded cause of death classified as diseases of the circulatory system. Secondary outcomes included prostate cancer–specific mortality (PCSM), defined as death attributed to prostate cancer, and all-cause mortality (ACM). For cause-specific mortality models, deaths from other causes was treated as competing events. Participants from New Mexico (n=69) were excluded from CVD mortality analyses due to missing cause-specific data.

### Covariates

Individual-level covariates included age at diagnosis, diagnosis year, race and ethnicity (non-Hispanic White, non-Hispanic Black, Hispanic, and Asian American/Native Hawaiian/Pacific Islander), insurance status (private, uninsured, Medicaid, Medicare, other), and marital status (married vs unmarried) and receipt of definitive local treatment (surgery or radiotherapy). Geospatial covariates were obtained at geomasked residence at diagnosis and included Census Tract-level neighborhood socioeconomic status and population density, residential greenness, and Census Region of residence (North, Midwest, South, West). Neighborhood socioeconomic status was derived from socioeconomic indicators following previously defined methods,^21^ and modeled in tertiles in adjusted survival models. Residential greenness was modeled using quintiles of satellite-derived Normalized Difference Vegetation Index (NDVI) within 270m of the participants’ address which has previously been linked to mortality in men with PCa.^22^ Population density was modeled as a binary variable using a threshold of 1,000 persons per square mile.

### Statistical analysis

Baseline characteristics were summarized overall and across PM2.5 quintiles. Continuous variables were summarized using medians and interquartile ranges, and categorical variables using frequencies and percentages.

Cox proportional hazards models were used to estimate associations between pollutant exposure and mortality outcomes. Proportional hazards were tested using Schoenfeld residuals, which showed no evidence of violations.^23^ For all-cause mortality, standard Cox models were fitted using time from diagnosis to death or censoring. For CVD-specific mortality and PCSM, cause-specific Cox models were fitted using stabilized inverse probability of censoring weights (IPCW) to account for competing deaths.^24^ IPCW were estimated using logistic regression models for death from competing causes conditional on baseline covariates, with stabilized weights trimmed at the 1st and 99th percentiles.

Models were fitted in sequential stages. The base model was stratified by age category and diagnosis year category. Fully adjusted models additionally included race and ethnicity, neighborhood socioeconomic status, insurance status, marital status, NDVI, population density, and census region. For continuous pollutant analyses, estimates were reported per IQR increase in exposure. For categorical pollutant analyses, hazard ratios were estimated for quintiles 2 through 5 relative to quintile 1. Tests for trend were performed by modeling the median pollutant concentration within each quintile as a continuous variable.

Missing covariate data were handled using multiple imputation with chained equations, generating five imputed datasets. Model estimates were pooled across imputed datasets using Rubin’s rules. Results are reported as hazard ratios (HRs) with 95% confidence intervals (CIs). All analyses were conducted using R.

## Results

### Study population

The final analytic cohort included 33,384 men with metastatic prostate cancer. Patients were evenly distributed across PM2.5 quintiles (Table 1). Baseline characteristics differed substantially across PM2.5 exposure groups. Patients in higher PM2.5 quintiles were slightly younger, more frequently Hispanic or non-Hispanic Black, more likely to reside in lower socioeconomic-status neighborhoods, and more likely to live in areas with higher population density and lower residential greenness. PM2.5 exposure was also strongly associated with calendar period and census region, with the highest PM2.5 quintile including a larger proportion of patients diagnosed in earlier years. Differences across PM2.5 quintiles were statistically significant for age, race and ethnicity, insurance status, diagnosis year, marital status, neighborhood socioeconomic status, census region, Gleason score, surgery, ADT, radiotherapy, population density, PM2.5, NO2, and NDVI (all p<0.001 except radiotherapy p=0.007; Table 1).

**Table 1.**
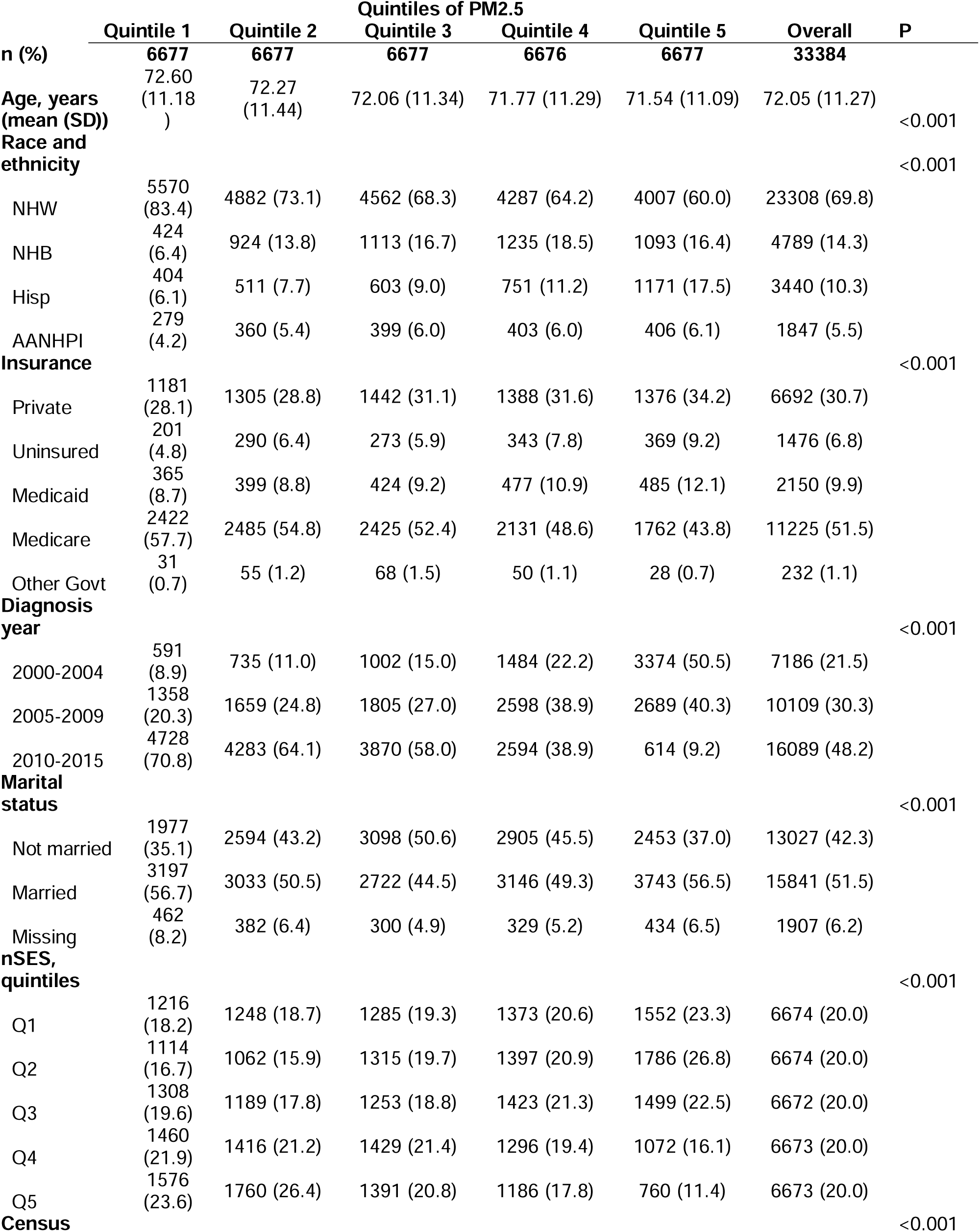
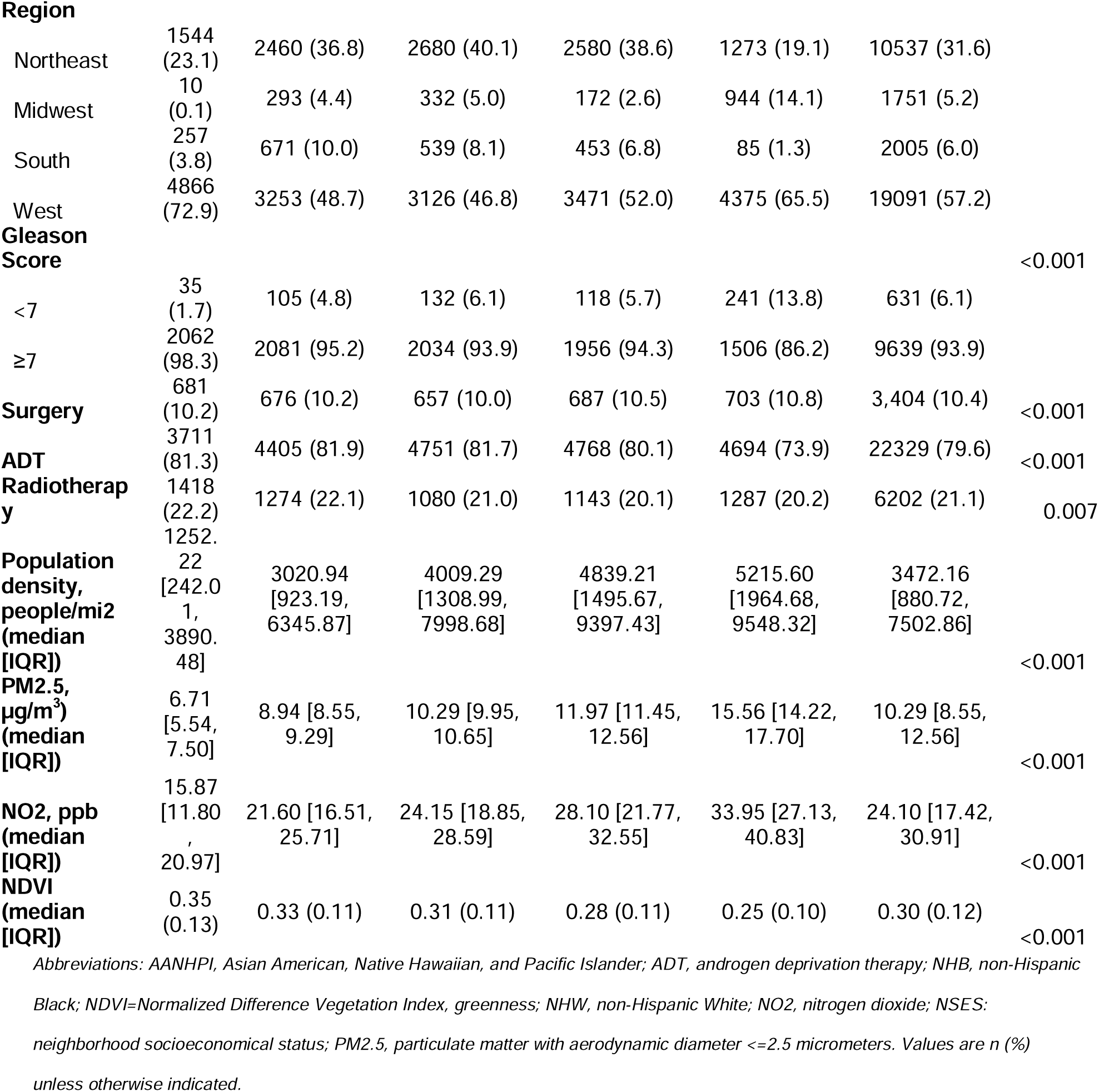
Baseline characteristics of men with metastatic prostate cancer, stratified by PM2.5 quintile.

### PM2.5, NO2, and mortality outcomes

In fully adjusted models, PM2.5 exposure was most consistently associated with cardiovascular mortality. Each interquartile range increase in PM2.5 was associated with higher cardiovascular mortality risk (HR 1.04, 95% CI 1.02–1.07), with evidence of a positive trend across PM2.5 quintiles (p-for-trend=0.008; Table 2). Compared with patients in the lowest PM2.5 quintile, those in the fourth and fifth quintiles had higher cardiovascular mortality risk (Q4: HR 1.18, 95% CI 1.01–1.37; Q5: HR 1.21, 95% CI 1.02–1.42; Table 2).

**Table 2.**
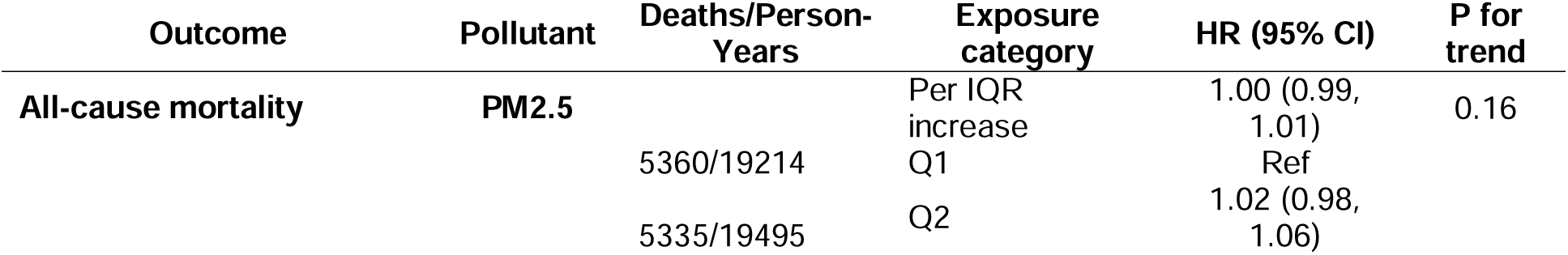
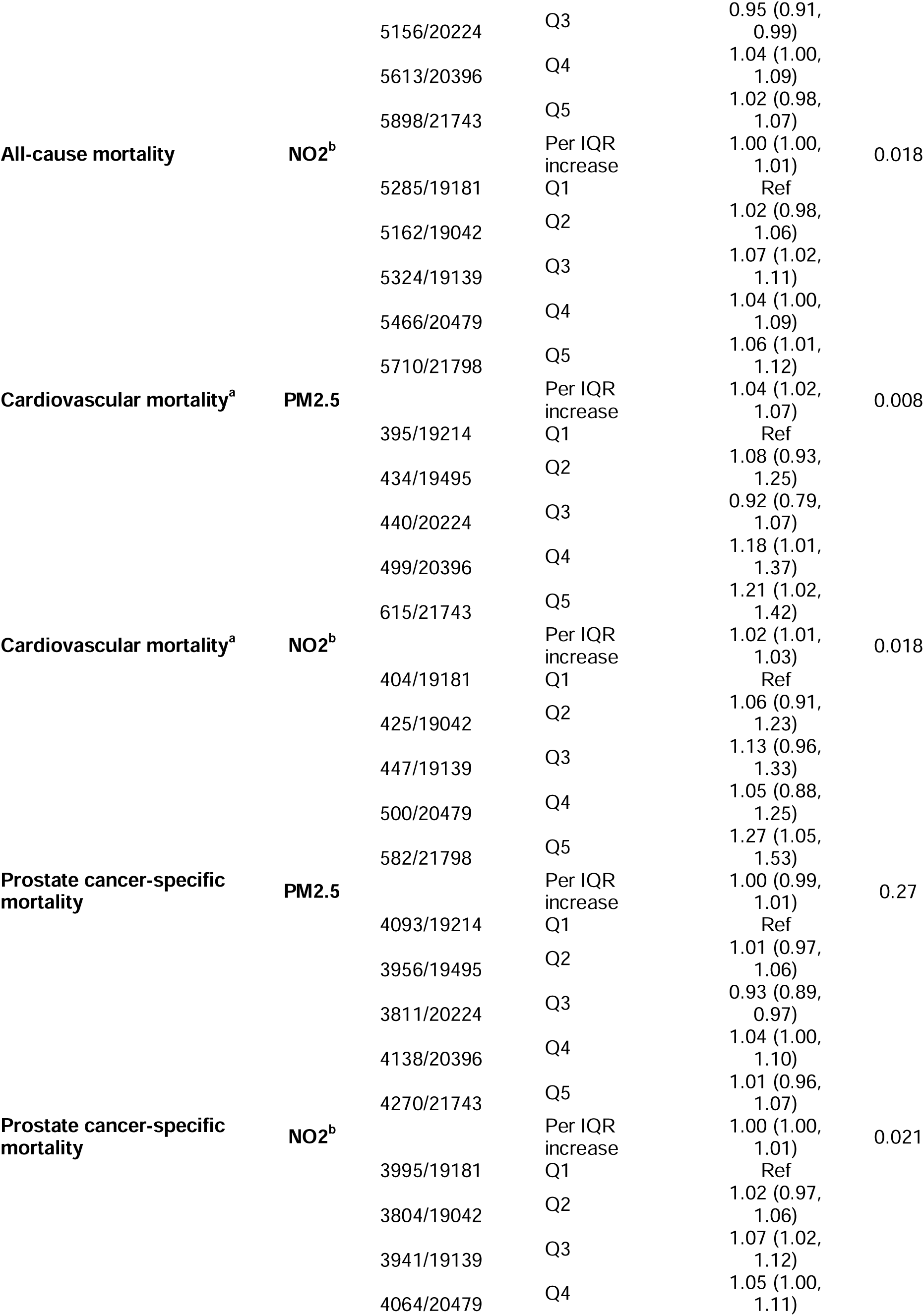
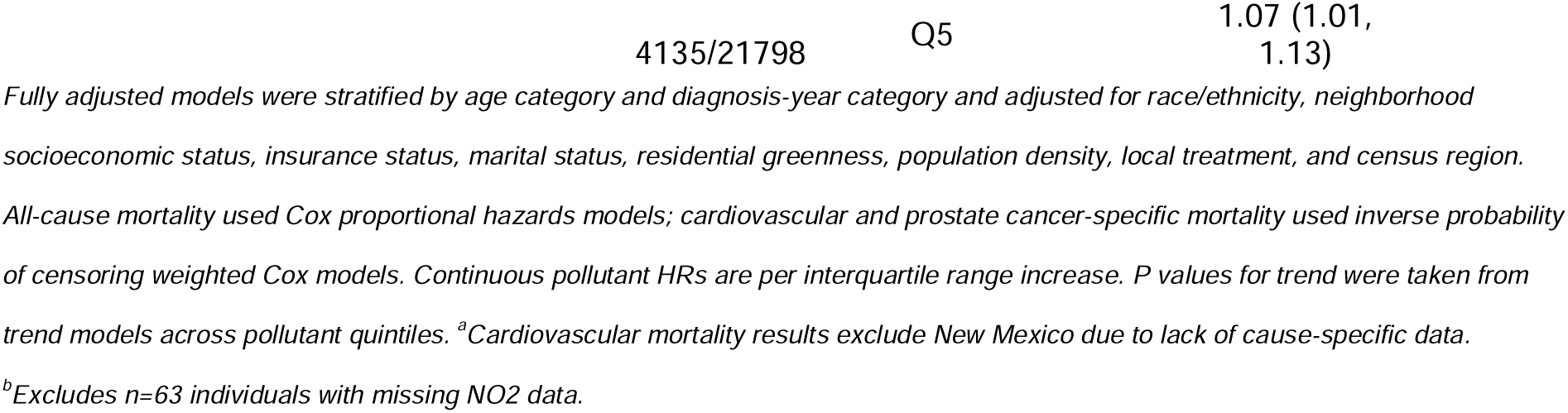
Fully adjusted associations of PM2.5 and NO2 with mortality outcomes.

Associations between PM2.5 and all-cause mortality were weaker. In the fully adjusted continuous model, PM2.5 was not associated with all-cause mortality (HR 1.00, 95% CI 0.99–1.01), and there was no clear trend across quintiles (p-for-trend=0.16; Table 2). Similarly, PM2.5 was not associated with prostate cancer-specific mortality when modeled continuously (HR 1.00, 95% CI 0.99–1.01), and the trend across quintiles was not statistically significant (p-for-trend=0.27; Table 2).

NO2 showed modest positive associations across mortality outcomes, although the pattern was less specific than for PM2.5. NO2 was associated with cardiovascular mortality when modeled continuously (HR 1.02, 95% CI 1.01–1.03), with a positive trend across quintiles (p-for-trend=0.02). Compared with the lowest NO2 quintile, the highest NO2 quintile was associated with higher cardiovascular mortality risk (Q5: HR 1.27, 95% CI 1.05–1.53; Table 2). Higher NO2 exposure was also associated with prostate cancer-specific mortality in quintile-based analyses (Q5 vs Q1: HR 1.07, 95% CI 1.01–1.13; p-for-trend=0.02). Associations with all-cause mortality were smaller, although the highest NO2 quintile was associated with modestly higher all-cause mortality compared with the lowest quintile (Q5: HR 1.06, 95% CI 1.01–1.12; p-for-trend=0.02; Table 2).

### Joint ADT–pollution exposure categories

Kaplan-Meier curves stratified by combined ADT–pollutant exposure groups demonstrated significant unadjusted differences in overall survival for both PM2.5 and NO2 categories (Figure 1). Because these curves were unadjusted, adjusted associations are reported in Table 3.

**Figure 1.**
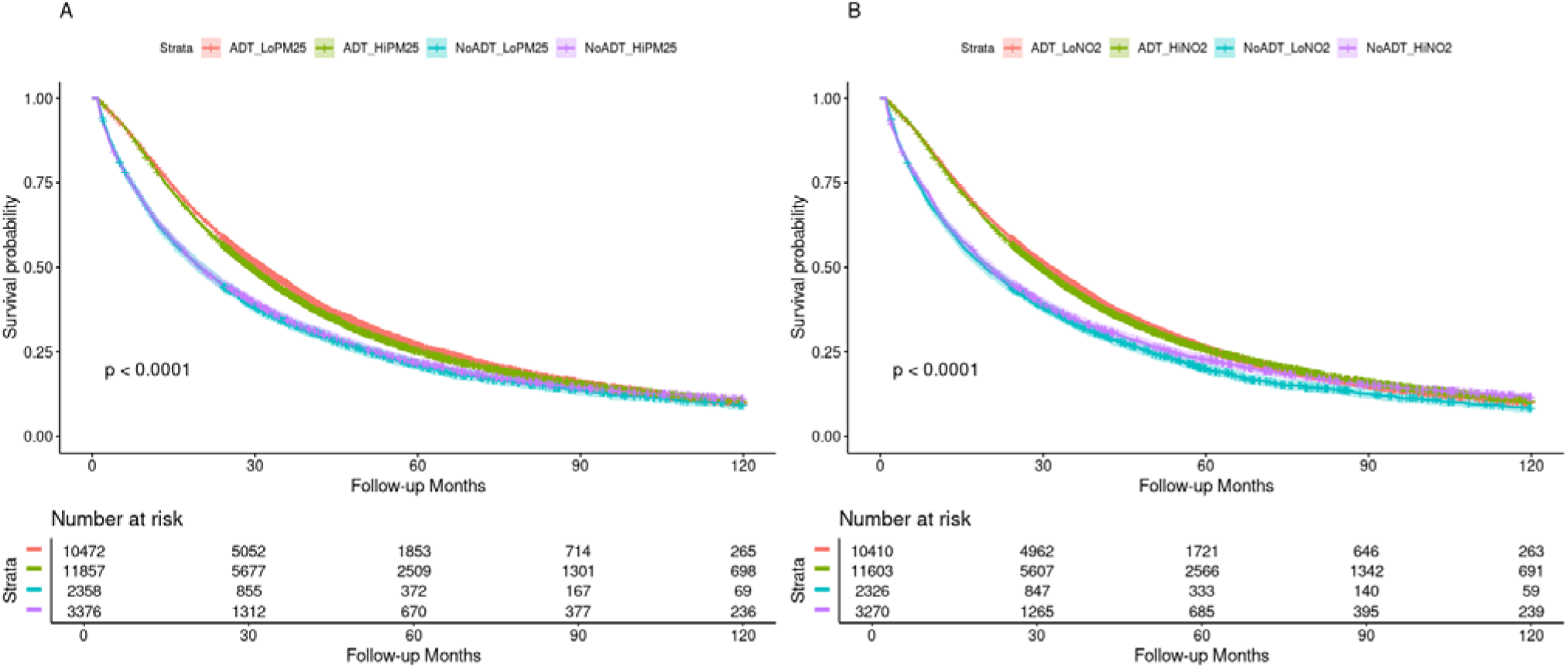
**Kaplan-Meier Survival Curves by Pollution and Androgen Deprivation Therapy Use in men with metastatic prostate cancer**

**Table 3.**
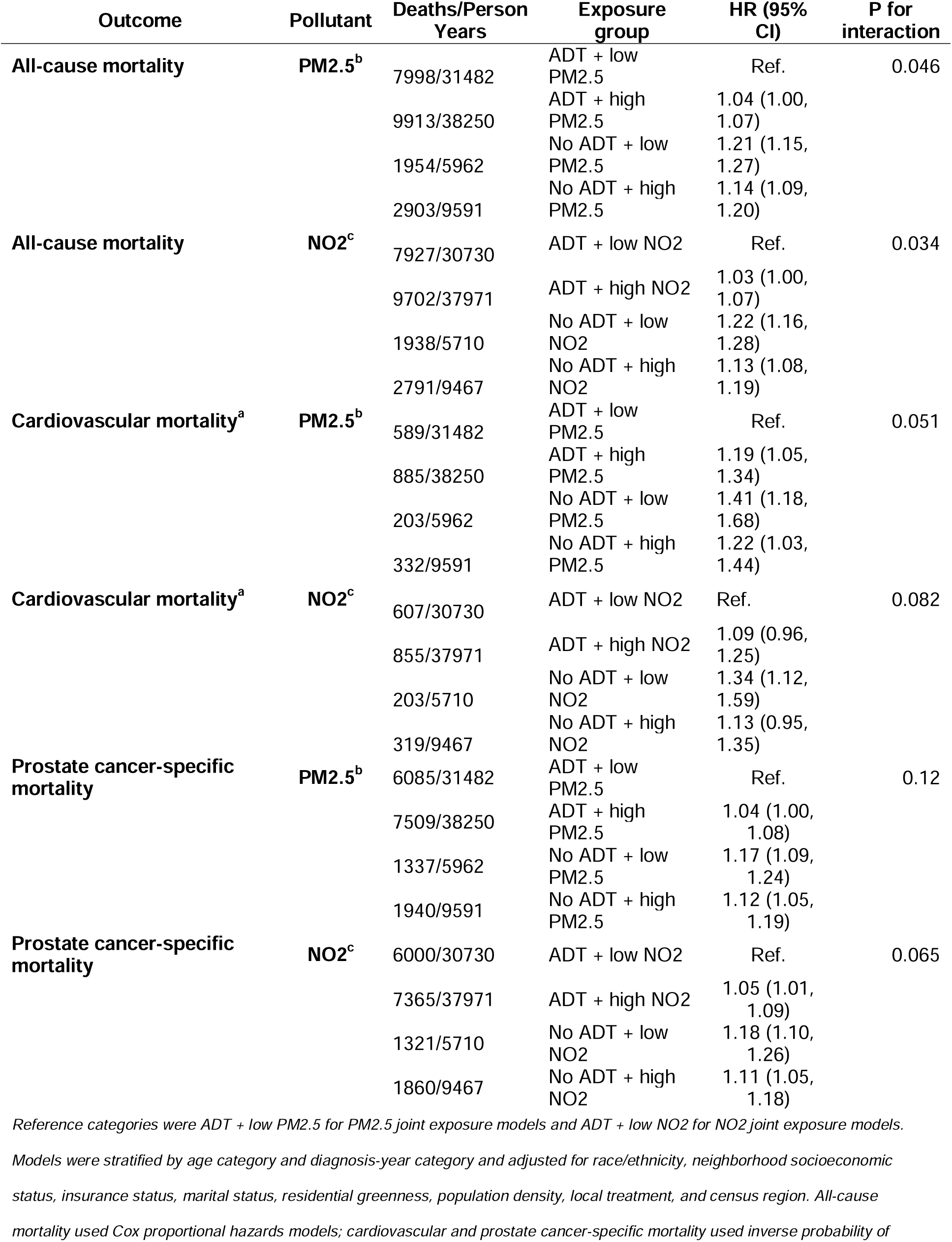
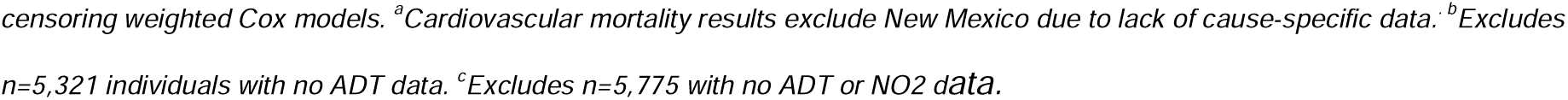
Fully adjusted joint ADT-pollution exposure categories and mortality outcomes.

| Outcome | Pollutant | Deaths/Person Years | Exposure group | HR (95% CI) | P for interaction |
| --- | --- | --- | --- | --- | --- |
| **All-cause mortality** | **PM2.5<sup>b</sup>** | 7998/31482 | ADT + low PM2.5 | Ref. | 0.046 |
|  |  | 9913/38250 | ADT + high PM2.5 | 1.04 (1.00, 1.07) |  |
|  |  | 1954/5962 | No ADT + low PM2.5 | 1.21 (1.15, 1.27) |  |
|  |  | 2903/9591 | No ADT + high PM2.5 | 1.14 (1.09, 1.20) |  |
| **All-cause mortality** | **NO2<sup>c</sup>** | 7927/30730 | ADT + low NO2 | Ref. | 0.034 |
|  |  | 9702/37971 | ADT + high NO2 | 1.03 (1.00, 1.07) |  |
|  |  | 1938/5710 | No ADT + low NO2 | 1.22 (1.16, 1.28) |  |
|  |  | 2791/9467 | No ADT + high NO2 | 1.13 (1.08, 1.19) |  |
| **Cardiovascular mortality<sup>a</sup>** | **PM2.5<sup>b</sup>** | 589/31482 | ADT + low PM2.5 | Ref. | 0.051 |
|  |  | 885/38250 | ADT + high PM2.5 | 1.19 (1.05, 1.34) |  |
|  |  | 203/5962 | No ADT + low PM2.5 | 1.41 (1.18, 1.68) |  |
|  |  | 332/9591 | No ADT + high PM2.5 | 1.22 (1.03, 1.44) |  |
| **Cardiovascular mortality<sup>a</sup>** | **NO2<sup>c</sup>** | 607/30730 | ADT + low NO2 | Ref. | 0.082 |
|  |  | 855/37971 | ADT + high NO2 | 1.09 (0.96, 1.25) |  |
|  |  | 203/5710 | No ADT + low NO2 | 1.34 (1.12, 1.59) |  |
|  |  | 319/9467 | No ADT + high NO2 | 1.13 (0.95, 1.35) |  |
| **Prostate cancer-specific mortality** | **PM2.5<sup>b</sup>** | 6085/31482 | ADT + low PM2.5 | Ref. | 0.12 |
|  |  | 7509/38250 | ADT + high PM2.5 | 1.04 (1.00, 1.08) |  |
|  |  | 1337/5962 | No ADT + low PM2.5 | 1.17 (1.09, 1.24) |  |
|  |  | 1940/9591 | No ADT + high PM2.5 | 1.12 (1.05, 1.19) |  |
| **Prostate cancer-specific mortality** | **NO2<sup>c</sup>** | 6000/30730 | ADT + low NO2 | Ref. | 0.065 |
|  |  | 7365/37971 | ADT + high NO2 | 1.05 (1.01, 1.09) |  |
|  |  | 1321/5710 | No ADT + low NO2 | 1.18 (1.10, 1.26) |  |
|  |  | 1860/9467 | No ADT + high NO2 | 1.11 (1.05, 1.18) |  |

Reference categories were ADT + low PM2.5 for PM2.5 joint exposure models and ADT + low NO2 for NO2 joint exposure models.
Models were stratified by age category and diagnosis-year category and adjusted for race/ethnicity, neighborhood socioeconomic status, insurance status, marital status, residential greenness, population density, local treatment, and census region. All-cause mortality used Cox proportional hazards models; cardiovascular and prostate cancer-specific mortality used inverse probability of
censoring weighted Cox models. <sup>a</sup>Cardiovascular mortality results exclude New Mexico due to lack of cause-specific data. <sup>b</sup>Excludes n=5,321 individuals with no ADT data. <sup>c</sup>Excludes n=5,775 with no ADT or NO2 data.

In fully adjusted joint ADT–PM2.5 models, among patients receiving ADT, high PM2.5 exposure was associated with an approximately 20% higher cardiovascular mortality compared with low PM2.5 exposure (HR 1.19, 95% CI 1.05–1.34; Table 3). This represented the strongest joint ADT–pollution signal. The interaction between ADT and PM2.5 for cardiovascular mortality was borderline (p-interaction=0.051). Among ADT-treated patients, high PM2.5 exposure was also associated with modestly higher all-cause mortality (HR 1.04, 95% CI 1.00–1.07) and prostate cancer-specific mortality (HR 1.04, 95% CI 1.00–1.08), although these associations were smaller.

In joint ADT–NO2 models, high NO2 exposure among ADT-treated patients was not clearly associated with cardiovascular mortality (HR 1.09, 95% CI 0.96–1.25), but was associated with prostate cancer-specific mortality (HR 1.05, 95% CI 1.01–1.09; Table 3). Patients without recorded ADT exposure had higher mortality across outcomes compared with the ADT with low pollutant exposure reference groups, regardless of pollutant exposure category. These comparisons should be interpreted cautiously because they reflect differences in both ADT status and pollution exposure and may be influenced by treatment selection, access to care, disease severity, or incomplete treatment information.

## Discussion

In this population-based cohort of men with metastatic prostate cancer, we found that fine particulate matter exposure was associated with cardiovascular mortality, but associations with all-cause mortality and prostate cancer-specific mortality were weaker. However, men with higher NO2 exposure had higher overall mortality and cause-specific mortality. In joint ADT-pollution analyses, ADT-treated patients exposed to higher PM2.5 had nearly 20% higher cardiovascular mortality compared to patients exposed to lower PM2.5. High versus low NO2 exposure among ADT-treated men was associated with higher mortality, but only the association with prostate cancer-specific mortality reached statistical significance. Taken together, these findings suggest that, among men with metastatic prostate cancer, the clearest signal relates to PM2.5 and cardiovascular mortality.

In the general population, ambient air pollution has been associated with cardiovascular morbidity and mortality.^25^ Among patients with cancer and cancer survivors, PM2.5 exposure has also been associated with higher cardiovascular and cardiopulmonary mortality, suggesting that cancer populations may represent a vulnerable group.^26^ We previously found in this cohort that higher exposure to PM2.5 components was associated with higher risks of all-cause and cardiovascular mortality.^10^ In addition, there is growing concern regarding stagnating progress in reducing nationwide PM2.5 exposure in light of the greater frequency and intensity of wildfires since 2016, particularly in Western states where declines have reversed.^13^ In another study using Surveillance, Epidemiology and

End Results (SEER) data from 2000 to 2021, higher wildfire-related PM2.5 was associated with higher cardiopulmonary and cancer mortality, with estimates as high as 15-25% elevated risk.^14^ Our study adds to this literature by focusing specifically on men with metastatic disease, in whom cardiovascular mortality remains clinically relevant and who may experience higher risks of CVD-related side effects of hormone therapy.^27,28^

The joint ADT-PM2.5 findings should be interpreted as hypothesis-generating. ADT is associated with adverse cardiometabolic effects and has been linked to cardiovascular events in prior studies.^4,5^ In our analysis, among men receiving ADT, higher PM2.5 exposure was associated with higher risk of cardiovascular death. This association alone is insufficient to establish that PM2.5 modifies the cardiovascular effects of ADT. However, it raises the possibility that environmental exposures may exacerbate the cardiovascular risk profile of patients receiving systemic therapy for metastatic prostate cancer. This interpretation aligns with the evolving cardio-oncology framework, in which cardiovascular outcomes among patients with cancer may be shaped by cancer therapy, baseline comorbidity, social conditions, and environmental exposures.^15,16^ The stronger association observed for cardiovascular mortality than for prostate cancer-specific mortality supports the interpretation that the observed signal is more likely related to cardiovascular vulnerability than to prostate cancer progression.

These findings may also be relevant from a public health “precision prevention” perspective that aims to tailor public health and clinical management of patients based on their individual and contextual factors that may increase susceptibility to certain diseases.^29^ In our cohort, higher PM2.5 exposure was more common among patients living in higher-density and more socioeconomically disadvantaged areas. This suggests that environmental exposure may overlap with other markers of social vulnerability. These data support further work evaluating whether environmental exposure can add contextual information to cardiovascular risk assessment in metastatic prostate cancer. More broadly, they reinforce the need to consider environmental conditions as part of the structural context in which cancer outcomes occur.^15,27,28,30^

Our study is not devoid of limitations. Pollutant exposure was assigned using residential address at diagnosis and may not capture prior exposure, residential mobility, or time spent away from home, but our model allowed finer scale linkages than earlier studies relying on ZIP code or county, and enabled linkage to over 30,000 individuals. ADT exposure was based on registry-recorded hormone therapy information and did not capture treatment duration, drug class, adherence, or subsequent treatment intensification. Residual miscoding or misclassification of cause of death is possible, particularly for cause-specific mortality but we expect this bias to be non-differential with respect to the exposure. Finally, residual confounding may persist, including by smoking, baseline cardiovascular disease, medication use, and other individual-level risk factors not fully captured in registry data.

In conclusion, higher PM2.5 exposure was associated with cardiovascular mortality among men with metastatic prostate cancer, whereas associations with all-cause mortality, prostate cancer-specific mortality, and NO2 were less consistent. Among ADT-treated men, higher PM2.5 exposure was also associated with higher hazards of cardiovascular death. These findings support further investigation of environmental exposures in cardiooncology, particularly as potential contributors to cardiovascular vulnerability among patients with advanced cancer.

## Data Availability

The data used in this study are not publicly available and cannot be shared under the terms of the data use agreement. Queries about the database and analytic code can be sent to the corresponding author.

